# Delayed-centrifugation blood collection tube for remote blood biomarker assessment

**DOI:** 10.64898/2026.10.30.26366958

**Authors:** You-Rim Lee, Anurag Raj, Oscar L. Lopez, Xuemei Zeng, Thomas K. Karikari

## Abstract

**Introduction:** Plasma phosphorylated tau 217 (p-tau217) and amyloid-β 42 (Aβ42) are advancing toward clinical use for Alzheimer’s disease (AD), but their reliability can be impacted by preanalytical handling. Although alternative collection tubes with distinct stabilization strategies—P100 (protease inhibitor) and HEMAcollect (hemolysis/platelet-activation minimization)—have been developed to stabilize plasma proteins, how well they preserve these markers remains unresolved; we therefore compared both against fresh standard EDTA tubes.

**Methods:** In 40 participants, plasma p-tau217 and Aβ42 were measured (Lumipulse) in paired fresh EDTA, P100, and HEMAcollect samples after 5-day ambient storage. We assessed concentration change, rank-order preservation (Spearman ρ), percentage difference, and diagnostic classification under the p-tau217/Aβ42 ratio and p-tau217 alone.

**Results:** P100 preserved p-tau217 favorably, with high rank-order correlation (ρ=0.98) and a modest median change (+12.1%), and its classification agreement with fresh EDTA reached 97.5% using p-tau217 alone. HEMAcollect showed greater p-tau217 signal loss (−57.5%). Aβ42 was reduced in both tubes (−72.8% P100; −95.5% HEMAcollect) with loss of rank order, so neither preserved the p-tau217/Aβ42 ratio.

**Discussion:** Stability was tube- and analyte-specific, and P100 consistently outperformed HEMAcollect—preserving p-tau217 rank order while HEMAcollect reduced signal across analytes. Although neither tube preserved the Aβ42-dependent ratio, P100 shows potential for p-tau217-based workflows. Preanalytical stability must be established analyte by analyte before alternative tubes enter clinical AD diagnosis.

## 1. Background

Blood-based biomarkers, particularly plasma phosphorylated tau 217 (p-tau217) and amyloid-β 42 (Aβ42), are advancing rapidly toward clinical use for detecting Alzheimer’s disease (AD) pathology[1, 2]. Their diagnostic reliability, however, depends on preanalytical conditions that are difficult to standardize outside specialized centers. Current protocols require prompt, temperature-controlled processing, because Aβ peptides in particular are inherently unstable and decline rapidly under delayed or ambient-temperature storage[3, 4]. These requirements limit biomarker testing in settings that lack immediate processing or cold-chain capacity.[5]

To relax these constraints, blood collection tubes with additive-based stabilization have been introduced, though they pursue fundamentally different strategies. The P100 tube (BD Biosciences) is spray-coated with a protease-inhibitor cocktail intended to suppress enzymatic degradation of labile peptides, and our group previously showed that it slows Aβ degradation relative to ethylenediaminetetraacetic acid (EDTA)[6]. More recently, the HEMAcollect•PROTEIN tube (DNA Genotek) was introduced with a proprietary stabilizing liquid that preserves draw-time plasma protein concentrations for up to seven days at ambient temperature by minimizing ex vivo hemolysis and platelet activation—that is, by limiting the release of cellular contents rather than by inhibiting proteolysis[7]. Whether these distinct mechanisms translate into comparable preservation of specific AD markers is unknown.

Critically, these tubes were developed primarily to preserve plasma proteins in general, and neither has been evaluated head-to-head for whether it preserves the low-abundance markers used for clinical AD classification. Both remain for research use only.

Here we directly compared P100 and HEMAcollect against paired fresh EDTA plasma—the current de facto reference—for p-tau217 and Aβ42 measured on the Lumipulse platform. Beyond concentration change, we asked whether each device preserves rank order and diagnostic classification, and whether any degradation is uniform across analytes or analyte-specific.

## 2. Method

### 2.1. Participants and sample preparation

Participants were recruited from the University of Pittsburgh Alzheimer’s Disease Research Center (ADRC). The Pitt-ADRC study was approved by the University of Pittsburgh Institutional Review Board (STUDY19110245), and all participants provided written informed consent. Venous blood was drawn sequentially into 10 mL EDTA (BD Biosciences, Franklin Lakes, NJ, USA; Cat. No. 366643), 8.5 mL P100 (BD Biosciences; Cat. No. 366448), and 9 mL HEMAcollect (DNA Genotek, Ontario, Canada; Cat. No. HCPP-RUO) tubes, in that order. Fresh EDTA tubes were centrifuged at 2,000 × g for 10 min at 4 °C to obtain plasma, which was immediately stored at −80 °C. The paired P100 and HEMAcollect tubes were left at ambient laboratory conditions for 5 days (20.4 ± 1.0 °C) before being centrifuged according to each manufacturer’s protocol and stored at −80 °C. The cohort comprised 40 participants (median age 76 years [interquartile range {IQR} 72–81]; 62% female; 39% apolipoprotein E (*APOE* ε4) carriers [15/38 with genotype available]; 21 cognitively unimpaired, 10 mild cognitive impairment, 8 Alzheimer’s disease, and 1 frontotemporal dementia); full demographic and clinical characteristics are provided in Table 1.

**Table 1.** Demographic and clinical characteristics of the study cohort.

| Number of participants |  | 40 |
|---|---|---|
| Sex | Male, n (%) | 15 (38) |
|  | Female, n (%) | 25 (62) |
| Age, years | median [IQR] | 76 [72–81] |
| Education, years | median [IQR] | 18 [16–18] |
| Self-identified racial identity | White, n (%) | 29 (72) |
|  | Black/African American, n (%) | 9 (22) |
|  | Asian Pacific, n (%) | 2 (5) |
| *APOE* ε4 carrier<sup>\*</sup> | n/known (%) | 15/38 (39) |
| Amyloid PET<sup>†</sup> | Positive, n/available (%) | 5/15 (33) |
|  | Negative, n/available (%) | 10/15 (67) |
| CDR, n | 0 | 12 |
|  | 0.5 | 21 |
|  | 1 | 3 |
|  | ≥2 | 4 |
| MMSE, n | Normal (≥24) | 14 |
|  | Mild (18–23) | 2 |
|  | Moderate (10–17) | 2 |
|  | Severe (<10) | 0 |
|  | Assessed, n‡ | 18 |
| Diagnosis grouping | Cognitively unimpaired, n | 21 |
|  | MCI, n | 10 |
|  | Alzheimer's disease, n | 8 |
|  | FTD, n | 1 |

<sup>\*</sup>APOE ε4 carriership was defined as the presence of at least one ε4 allele; percentages are among participants with available genotype (n/known). Genotype was unavailable for 2 participants.
<sup>†</sup>Amyloid PET (Centiloid) data were available for 15 of 40 participants; amyloid status is based on the Centiloid-derived positivity classification, and percentages are among participants with available status.
‡MMSE was assessed in 18 of 40 participants and summarized by standard severity bands (normal ≥24; mild 18–23; moderate 10–17; severe <10); given the limited number assessed, these counts should be interpreted with caution.
General notes: Continuous variables (age, education) are presented as median [IQR] given the small sample size and non-normal distributions. Age was calculated at the date of plasma collection. CDR is summarized as the number of participants at each global score (0, 0.5, 1, ≥2) and was available for all 40 participants. Values of 0 indicate that no participant fell in that category for a variable with complete data. Diagnosis grouping: AD (probable AD, including atypical/SWML variants); MCI (amnestic/non-amnestic, including focal); FTD (temporal-variant frontotemporal dementia); cognitively unimpaired (controls, normalized, subjective complaint with normal scores, or abnormal scores without complaint).
Abbreviations: AD, Alzheimer's disease; APOE, apolipoprotein E; CDR, Clinical Dementia Rating; FTD, frontotemporal dementia; IQR, interquartile range; MCI, mild cognitive impairment; MMSE, Mini-Mental State Examination; PET, positron emission tomography.

### 2.2. Biomarker measurement

Plasma p-tau217 and Aβ42 were measured on a Lumipulse G1200 analyzer (Fujirebio) using marker-specific in vitro diagnostic (IVD) kits, following the manufacturer’s recommended protocol. The limits of detection and quantification were 0.041 and 0.047 pg/mL for p-tau217, and 0.207 and 0.8 pg/mL for Aβ42, respectively. All samples were analyzed on the same day. Calibration curves were generated using the manufacturer’s calibrator kit, and control samples from the sample control kit were run alongside study samples. Calibration curves showed R^2^ ≥ 0.99, and all control samples fell within the manufacturer-supplied acceptable concentration ranges.

### 2.3. Diagnostic classification

Diagnostic classification obtained from fresh EDTA plasma—the current standard sample type in clinical practice—was set as the within-participant reference, against which P100 and HEMAcollect results were compared. Classification was evaluated under three schemes. First, the p-tau217/Aβ42 ratio was classified using U.S. Food & Drug Administration (FDA) approved IVD diagnostic cutoffs, categorizing samples as Negative (≤0.00370; unlikely to have amyloid pathology), Indeterminate (0.00371–0.00737), or Positive (≥0.00738; likely to have amyloid pathology)[8]. Second, a data-driven cutoff for the p-tau217/Aβ42 ratio was derived from fresh EDTA values in the present Lumipulse dataset by receiver operating characteristic (ROC) analysis, yielding a single threshold (>0.007860) that defined a two-category Positive/Negative split. Third, p-tau217 alone was classified using cutoffs reported in a previous study (Negative <0.22, Indeterminate 0.22–0.34, Positive >0.34 pg/mL)[1].

### 2.4. Statistical analysis

Statistical analyses were performed using R version 4.6.0. Distributional normality was assessed with the Shapiro–Wilk test; given non-normal distributions, nonparametric methods were used. Paired comparisons between each stored tube (P100 or HEMAcollect) and fresh EDTA were tested with the Wilcoxon signed-rank test. Rank-order preservation between stored and reference measurements was assessed using Spearman correlation coefficients for each biomarker.

Percentage difference relative to fresh EDTA was calculated for each participant as (stored − EDTA) / EDTA × 100, where “stored” denotes the P100 or HEMAcollect measurement. ROC curve analysis, with the optimal cutoff determined by Youden’s index, was used to derive the data-driven ratio threshold. A significance level of *p* < 0.05 was used for all tests.

## 3. Results

### 3.1. P100 preserves p-tau217 but not Aβ42

To assess stability, we compared concentration, rank-order correlation, and percentage difference relative to EDTA for each marker and tube. In P100, both plasma p-tau217 and Aβ42 showed a significant decrease in concentration (p < 0.001; Figure 1A, 1C). However, p-tau217 remained highly correlated with EDTA, preserving rank order (ρ = 0.98, p < 0.001; Figure 1B), and the median difference relative to EDTA was only modest (+12.05%). In contrast, Aβ42 did not preserve rank order (ρ = 0.40, p = 0.011; Figure 1D) and decreased substantially across all participant samples, with a median difference of −72.80%. Thus, in P100, p-tau217 was preserved at the rank level, however Aβ42 was not.

**Figure 1.**
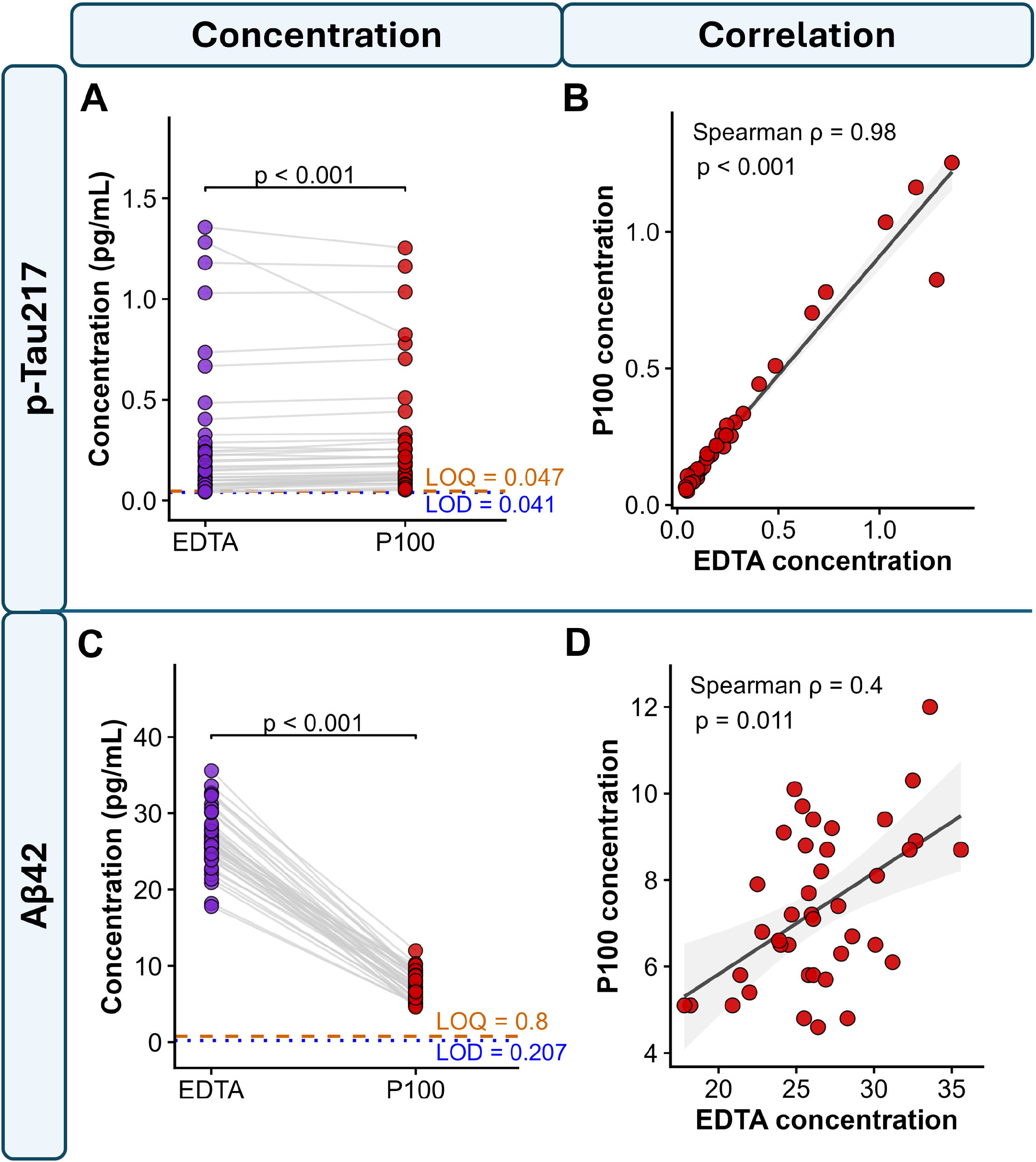
Stability of p-tau217 and Aβ42 in P100. Paired comparison of plasma p-tau217 (A–B) and Aβ42 (C–D) between fresh EDTA (reference) and P100 after 5-day ambient storage (n=40 participants). (A, C) Concentrations in EDTA versus P100; connecting lines indicate paired samples, and dashed lines denote the assay limit of detection (LOD) and limit of quantification (LOQ) (p-tau217: LOD=0.041, LOQ=0.047 pg/mL; Aβ42: LOD=0.207, LOQ=0.8 pg/mL). (B, D) Rank-order correlation between EDTA and P100 (Spearman ρ). P values are from Wilcoxon signed-rank tests. Abbreviations: Aβ42, amyloid-β 42; EDTA, ethylenediaminetetraacetic acid; LOD, limit of detection; LOQ, limit of quantification; p-tau217, phosphorylated tau 217

### 3.2. HEMAcollect reduces both p-tau217 and Aβ42

Stability in HEMAcollect was assessed in the same manner as for P100. Both p-tau217 and Aβ42 showed a significant decrease in concentration (p < 0.001; Figure 2A, 2C). As with P100, p-tau217 remained highly correlated with EDTA and preserved rank order (ρ = 0.95, p < 0.001; Figure 2B). However, its median percentage difference reached −57.54%, indicating substantial signal loss and a greater change than in P100. For Aβ42, the correlation with EDTA was even lower than in P100 and non-significant (ρ = 0.25, p = 0.124; Figure 2D), and the median percentage difference approached the assay floor at −95.53%. Thus, HEMAcollect reduced p-tau217 markedly beyond the loss seen in P100, and Aβ42 was lost almost entirely.

**Figure 2.**
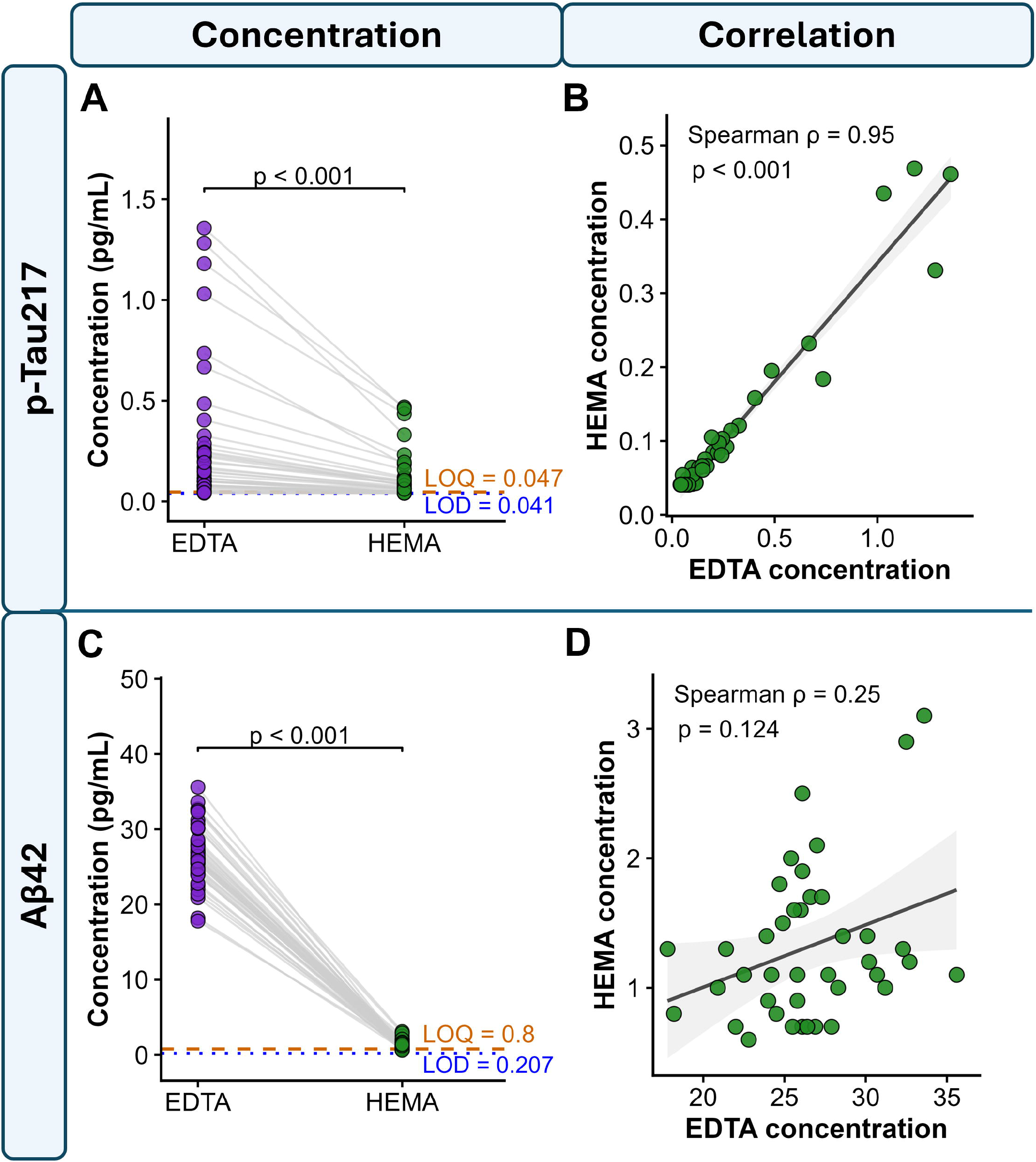
Stability of p-tau217 and Aβ42 in HEMAcollect. Paired comparison of plasma p-tau217 (A–B) and Aβ42 (C–D) between fresh EDTA (reference) and HEMAcollect after 5-day ambient storage (n=40 participants). Panels, symbols, assay LOD/LOQ lines, and statistical tests are as in Figure 1, with HEMAcollect replacing P100. Abbreviations: Aβ42, amyloid-β 42; EDTA, ethylenediaminetetraacetic acid; LOD, limit of detection; LOQ, limit of quantification; p-tau217, phosphorylated tau 217.

### 3.3. Diagnostic classification using the FDA-cleared p-tau217/Aβ42 ratio vs. p-tau217 alone

To determine whether diagnostic performance changed relative to EDTA in each tube, we evaluated classification using three approaches: the ratio-based IVD cutoffs (as our Lumipulse measurements used IVD kits), ratio-based cutoffs derived from a ROC analysis of the EDTA D0 concentration data itself, and, as an exploratory analysis, p-tau217 alone using cutoffs from a recently published reference. Under the ratio, agreement with the reference was 45% (IVD) / 50% (data-driven) for P100 and 37.5% / 37.5% for HEMAcollect, with all HEMAcollect samples classified Positive—a false-positive drift arising from Aβ42 reduction inflating the ratio. Using p-tau217 alone, agreement was 97.5% for P100 and 70% for HEMAcollect (12/40 reclassified, all toward Negative), reflecting a false-negative drift. Thus, the same tube produced diagnostic errors in opposite directions depending on the analyte combination used, and P100 markedly outperformed HEMAcollect under every scheme (Table 2).

**Table 2.**
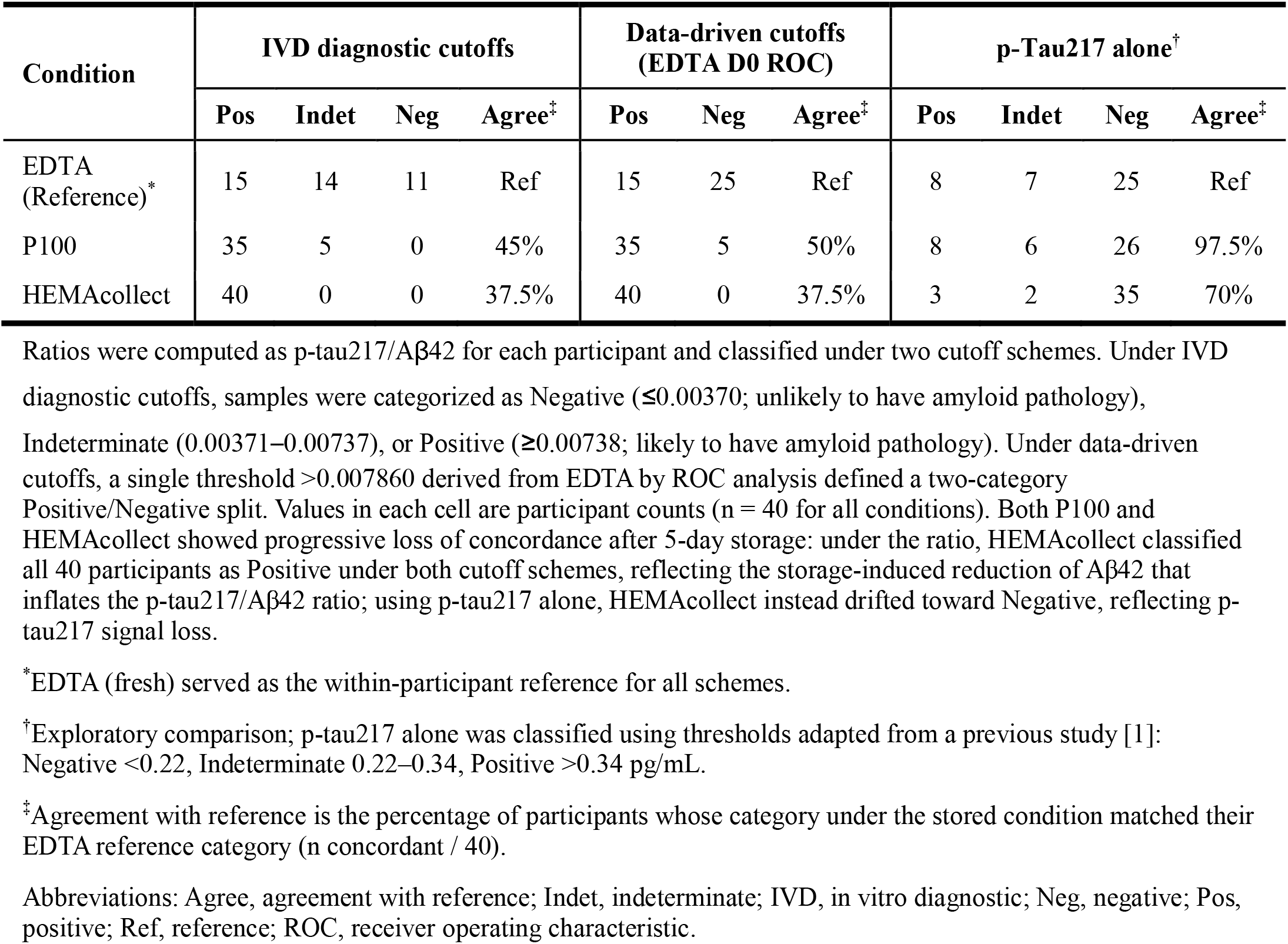
Diagnostic classification by the p-tau217/Aβ42 ratio and p-tau217 alone (n=40).

## 4. Discussion

In this paired, within-participant comparison of P100 and HEMAcollect against fresh EDTA, biomarker stability was strongly analyte- and tube-specific, and the two tubes were not equivalent: P100 preserved p-tau217 well enough such that the original diagnostic category of participants measured under pristine conditions was retained for nearly all (39/40) participants, whereas HEMAcollect reduced signal across all analytes and appeared unsuitable under every readout examined.

Plasma Aβ42 levels were significantly reduced in both P100 and HEMAcollect; more severely in HEMAcollect. Aβ peptides are inherently unstable and decline under ambient storage[3, 4], so the reduction itself is consistent with prior reports rather than a novel finding. Although the exact additive composition of HEMAcollect is not disclosed, its formulation offered no advantage over P100 for Aβ42 preservation. This is consistent with its stabilization mechanism, which targets ex vivo hemolysis and platelet activation rather than the proteolytic pathways that drive Aβ degradation; minimizing cellular contamination does not protect an already labile peptide from enzymatic cleavage.

Plasma p-tau217 also declined, but in P100 it retained a high correlation with EDTA (preserved rank order) and a stable median difference of only +12.05%. Because rank order was preserved even as absolute concentrations shifted, a tube-specific recalibration remains feasible for p-tau217 in P100—a shift in level can be corrected when relative ordering is retained, whereas it cannot once ordering is lost. The protease-inhibitor content of the P100 tube may contribute to this preservation of labile peptides—consistent with our group’s previous finding that P100 slowed Aβ degradation relative to EDTA[6]—although the specific inhibitors responsible were not examined here.

One point warranting consideration is the opposite direction of diagnostic error between the two readouts. The ratio drifted toward false-positive classification and p-tau217 alone toward false-negative—each a downstream consequence of the analyte-specific behavior described above (Aβ42 reduction inflating the ratio; p-tau217 signal loss lowering standalone values). When analyzing samples exposed to ambient storage, the stability of the marker of interest must therefore be defined for the specific analyte and readout combination, a consideration that future studies should incorporate.

Our findings extend, but also appear to diverge from, our group’s earlier report that P100 better preserved Aβ relative to EDTA[6]. Several differences likely account for this. That study assessed shorter room-temperature intervals (up to 24–96 h) and measured Aβ40 and Aβ42 by immunoprecipitation–mass spectrometry and Simoa, whereas the present study used a longer 5-day ambient storage and measured Aβ42 on the Lumipulse platform. The protective effect of P100 on Aβ that is detectable over hours may thus be overwhelmed by the more severe degradation that occurs over several days, and platform differences in the measured analyte may further contribute. In other words, P100 remains the better of the two tubes for Aβ42, but its advantage is insufficient to preserve diagnostically usable Aβ42 signal under the storage conditions tested here.

This study has several limitations. We evaluated a single storage condition—5 days at ambient temperature, chosen to provide a reasonable time window for transferring samples from real-world field work to a laboratory for assessment—and validation across a broader range of storage durations and temperatures will be needed to define the preanalytical window for each tube. The diagnostic markers were measured on a single platform (Lumipulse), so cross-platform confirmation is warranted. Aβ40 was not measured (restricting us to the p-tau217/Aβ42 ratio), the p-tau217 cutoffs were exploratory and externally derived, and the reference standard for the ROC analysis was limited. Finally, both tubes are currently for research use only, and any diagnostic application would require dedicated validation.

Nonetheless, P100 showed favorable preservation of p-tau217 over five days of ambient storage, suggesting potential for its future use in p-tau217-based workflows, whereas neither tube adequately preserved the Aβ42-dependent ratio. To our knowledge, this is the first evaluation of the HEMAcollect tube for clinical AD biomarkers. More broadly, preanalytical factors from blood collection through biomarker measurement are decisive and vary from one analyte to another; these analyte-specific effects must be characterized in advance so that stability is established before alternative tubes can be translated into clinical practice.

## Data Availability

The data supporting this study contain sensitive human participant information and cannot be deposited in a public repository due to institutional review board restrictions and the terms of participant consent. Specific de-identified data can be shared with qualified and identifiable investigators for the purpose of replicating the results and procedures in the study. Requests will be reviewed to ensure that data sharing requests conform to prevailing US legislation on data protection, intellectual property and confidentiality obligations. Establishment of a data transfer agreement may be necessary.

## 8. Acknowledgments

None

## 9. Conflicts of interest

X.Z. and T.K.K. are inventors on University of Pittsburgh patents and provisional patents regarding biofluid biomarker methods, targets and reagents/compositions, that may generate income for the institution and/or self should they be licensed and/or transferred to other organization(s). T.K.K. has served as an ad hoc consultant and/or advisory board member for Quanterix Corporation, SpearBio Inc., Alzheon, Siemens Healthineers and Neurogen Biomarking LLC., outside the submitted work. T.K.K. has received royalties from Bioventix for the transfer of specific antibodies and blood biomarker assays to third party organizations. The other authors have nothing to declare.

## 10. Funding sources

Thomas K. Karikari and the Karikari laboratory were supported by NIH/NIA (U01 AG098641, R01 AG083874, U24 AG082930, P30 AG066468, RF1 AG077474, R01 AG083156, R37 AG023651, R01 AG025516, R01 AG073267, R01 AG075336, R01 AG074971, U19 AG073153, U01 AG079828, P01 AG025204), NIH/NINDS (U01 NS131740, U01 NS141777, U24 NS141780), NIH/NIMH (R01 MH108509, R01 MH135096), Aging Mind Foundation (DAF2255207), DoD (HT94252320064), American Heart Association (24SFRNPCN1280228),

Wellcome Leap (CARE-2025-7270262472), the Chuck Noll Foundation for Brain Injury Research, Greater Houston Community Foundation, Fujirebio, Genentech, and Astellas Pharma US. The content of this article is solely the responsibility of the authors and does not necessarily represent the official views of the funders.

## 11. Consent statement

All participants provided written informed consent.

## Notes

### Author Declarations

The Pitt-ADRC study was approved by the University of Pittsburgh Institutional Review Board (STUDY19110245), and all participants provided written informed consent.

